# A multivalent RSV vaccine based on the Modified Vaccinia Ankara vector shows moderate protection against disease caused by RSV in older adults in a Phase 3 clinical study

**DOI:** 10.1101/2024.07.22.24309971

**Authors:** Elke Jordan, Victoria Jenkins, Günter Silbernagl, Maria Paulina Velasco Chávez, Darja Schmidt, Frauke Schnorfeil, Stephanie Schultz, Liddy Chen, Fernanda Salgado, Jeanne-Marie Jacquet, Tobias Welte, Laurence De Moerlooze

## Abstract

Respiratory syncytial virus (RSV) causes a significant disease burden in older adults. The live recombinant vaccine based on a nonreplicating modified vaccinia Ankara (MVA-BN) poxvirus, MVA-BN-RSV, encoding for multiple proteins of RSV subtypes A and B, was assessed for efficacy against respiratory disease caused by RSV.

Adults aged ≥ 60 years, with or without underlying chronic conditions, were enrolled and randomized in a 1:1 ratio to receive a single dose of vaccine or placebo and were followed for disease caused by RSV infection during the 2022-2023 season. The 2 primary endpoints were RSV-associated lower respiratory tract disease (LRTD) with ≥ 3 and ≥ 2 symptoms; acute respiratory disease (ARD) was a key secondary endpoint. The humoral RSV-specific immune response was assessed at baseline and 14 days post-vaccination. Safety was evaluated by collection of solicited adverse events (AEs) and unsolicited AEs for 7 and 28 days post-vaccination respectively, and SAEs for the entire study period.

In total, 18348 participants were included in the final efficacy and safety analyses. Vaccine efficacy was 42.9% (95% CI: -16.1; 71.9) against RSV-associated LRTD with ≥ 3 symptoms, 59.0% (95% CI: 34.7; 74.3) against LRTD with ≥ 2 symptoms, and 48.8% (95% CI: 25.8; 64.7) against ARD. The primary objective was not met for LRTD with ≥ 3 symptoms since the lower bound of the 95% CI was below 20%, the prespecified success criterion. The vaccine-elicited immune response showed mean fold-increases of 1.7 for RSV A and B neutralizing antibodies and 2.9 and 4.3 for RSV-specific IgG and IgA, respectively. The vaccine displayed mild to moderate reactogenicity, and no safety concerns were identified.

MVA-BN-RSV induced suboptimal protection against RSV-associated LRTD, likely due to suboptimal neutralizing antibody response. The vaccine had an acceptable safety profile and confirmed immunogenicity, overall showing promise for MVA-BN-vectored constructs targeting other diseases.

## 1. Introduction

Respiratory Syncytial Virus (RSV) is a cause of severe respiratory disease in children below the age of 5 years, where the highest disease burden is observed, but also in older adults, and adults with underlying comorbidities [1]. Meta analyses estimate the yearly global incidence of RSV-acute respiratory infection (ARI) to be 6.7 per 1000 adults ≥ 65 years of age worldwide [2], and 16.2 per 1000 adults ≥ 60 years of age in high income countries [3]. The associated estimated hospitalization rates are 1.0 per 1000 and 1.5 per 1000, respectively [2, 3].

In the long search for an RSV vaccine, the F protein was recognized as an important antigen able to elicit a protective humoral immune response, and the preF conformation of the protein was identified as the target of naturally induced antibodies during RSV infection [4]. Recent RSV vaccine candidates have used the preF protein as the vaccine antigen in varied approaches: protein, mRNA or vector based [5]. Two vaccines became available in 2023 for the immunization of adults from the age of 60 years [6, 7]; both are protein-based and use the RSV F protein, stabilized in its preF conformation. These vaccines elicit a high neutralizing antibody response and have shown a protective efficacy for one RSV season of more than 80% in adults ≥ 60 years against RSV-associated lower respiratory tract disease (LRTD) [8, 9] and up to 94.1% against severe disease [8]. Data available beyond the initial analysis, for a longer follow-up, reveal variable durability of efficacy according to the platform. A drop in efficacy against RSV-associated LRTD was already visible from 82.4% to 63.0% from 3.7 to 8.6 months of follow-up in one RSV season for an mRNA vaccine [10, 11]; a decrease in efficacy was visible in the second RSV season for the available protein-based vaccines [12, 13] with an approximate drop of -10% to -30%,depending on the study and the LRTD endpoint; while for an adenovirus-vectored vaccine, sustained protection against RSV-associated LRTD was observed in a third season of follow-up in phase 2 (78.7% in season 3 versus 80.% in season 1) [14, 15], and no evidence of a decline in a second season compared to season 1 was apparent in phase 3 [16].

The MVA-BN-RSV vaccine candidate was developed as an alternative, multiple antigen approach, using the modified vaccinia Ankara (MVA-BN) poxvirus as a vector to express 5 RSV proteins: the F protein, which is expressed both in the preF and postF conformations, the G protein of RSV types A and B, and the internal M2-1 and N proteins [15]. The potential advantages of a multi-component vaccine, i.e. the induction of CD4+ and CD8+ T cell responses, along with antibodies including IgAs, were demonstrated in preclinical studies [17]. The immunogenicity profile of MVA-BN-RSV was characterized in Phase 1 and 2 clinical studies. The vaccine elicited RSV-specific IgG and IgA and a neutralizing response against both RSV A and B subtypes [18, 19]. This was accompanied by broad T cell responses as measured against 5 peptide pools representing the 5 encoded antigens [18, 19]; fold-increases in T cell responses appeared higher than the F-specific response reported for preF focused vaccines [16]. This distinctive balance of humoral responses and strong cellular responses stimulated by multiple antigens was thought to contribute to different pathways of protection [16] and suggested potential for a multi-faceted mode of action. MVA-BN-RSV was subsequently shown to prevent RSV colonization in an RSV challenge trial [20]. The Phase 3 study reported here aimed to assess the protective efficacy of the vaccine against LRTD associated with RSV in adults ≥ 60 years of age.

## 2. Methods

### 2.1. Study design, population, and vaccination

This was a randomized, double-blind, multicenter, placebo-controlled phase 3 study in adults ≥ 60 years of age, which was conducted at over 100 sites in the US and Germany between April 2022 and September 2023. Independent ethics committees or institutional review boards for each trial site approved the study protocol and its amendments.

All study participants were apprised of the trial aspects and signed an informed consent before any study procedure. The study was conducted according to the ethical principles of the Declaration of Helsinki [21], and Good Clinical Practice guidelines [22] and the applicable regulatory requirements. Safety during the trial was monitored by an Independent Data Monitoring Committee, which reviewed unblinded safety data at regular intervals.

Study participants were randomly assigned in a 1:1 ratio to 2 groups to receive a single dose of MVA-BN-RSV or placebo, administered intramuscularly in the deltoid muscle.

Participants could have one or more, clinically stable, chronic medical conditions such as chronic cardiac disease and chronic lung disease, congestive heart failure, hypertension, type 2 diabetes mellitus, hyperlipoproteinemia, or hypothyroidism. Frailty was assessed for each participant using the Tilburg frailty indicator. Previous vaccination with an RSV vaccine, or any planned vaccination with an RSV vaccine other than the trial vaccine was an exclusion criterion.

MVA-BN-RSV consists of MVA-BN [23] encoding 5 codon optimized RSV antigens: F (subtype A, strain A long), G (subtype A, strain A2), G (subtype B, strain 185 37), N, and M2-1 (both strain A2). MVA-BN-RSV and placebo were supplied by Bavarian Nordic A/S, Denmark. MVA-BN-RSV was provided as a liquid frozen formulation containing at least 3 × 10^8^ infectious units per 0.5 mL and placebo consisted of Tris-buffered saline.

### 2.2. Objectives

The primary objective of the study was to assess the clinical efficacy of the MVA-BN-RSV vaccine against LRTD associated with RSV in adults ≥ 60 years of age, during one RSV season. Two primary endpoints were defined: RSV-associated LRTD with at least 3 symptoms and RSV-associated LRTD with at least 2 symptoms; vaccine efficacy (VE) was assessed for endpoints reported from 14 days post-vaccination until the end of one RSV season. A key secondary objective was to assess VE against RSV-associated ARD during one RSV season. The assessment of VE against severe LRTD was also a secondary objective, and the assessment of VE against hospitalization or mortality related to confirmed RSV disease was an exploratory objective. A supplementary analysis of VE according to RSV subtype was performed, and subgroup analyses of efficacy according to age, sex, race, frailty, pre-existing chronic conditions and region were conducted. Reactogenicity, safety and immunogenicity were assessed as secondary objectives.

### 2.3. Efficacy assessments

Participants were followed for disease surveillance via monthly telephone calls and electronic alerts 3 times a week until the end of the RSV season. The occurrence of respiratory tract symptoms led to a visit at the study site to confirm symptoms and to collect a nasopharyngeal swab within 5 days of symptom onset for polymerase chain reaction (PCR) testing at the central laboratory.

RSV-associated ARD was defined by the presence of either 1 ARD symptom lasting for at least 24 hours or 2 simultaneously occurring ARD symptoms (irrespective of duration), with onset at least 14 days following vaccination until the end of one RSV season, with RSV- association confirmed by PCR. ARD symptoms included: rhinorrhea, nasal congestion, pharyngitis, earache, new cough or worsening of pre-existing cough, new wheezing or worsening of pre-existing wheezing, new sputum production or worsening of pre-existing sputum production, new shortness of breath or worsening of pre-existing shortness of breath, fever > 100°F / > 37.8°C (oral temperature). RSV-associated LRTD was defined as i) severe LRTD, i.e. the presence of at least 1 of the following signs: hypoxemia (oxygen saturation < 92% at rest in conjunction with an at least 3% decrease from baseline), respiratory rate > 25 breaths/Min, imaging evidence of new onset of bronchitis, bronchiolitis, or pneumonia, or ii) LRTD with at least 3 or at least 2 of the following symptoms: new wheezing or worsening of pre-existing wheezing, new shortness of breath or worsening of pre-existing shortness of breath, new cough or worsening of pre-existing cough, new sputum production or worsening of pre-existing sputum productions, signs of severe LRTD. All documented respiratory tract symptoms with PCR-confirmed RSV positivity were adjudicated to be either ARD, LRTD (severe, with 3-symptoms, or with 2-symptoms) or no endpoint by an independent adjudication committee.

Study participants were also asked to complete the Respiratory Intensity and Impact Questionnaire (RiiQ) [24] to assess changes in symptoms after RSV symptom onset until symptoms resolved or until 21 days. Symptoms were self-reported based on a grading scale: none, mild, moderate, severe.

### 2.4. Immunogenicity assessments

Serum antibody responses were assessed in a subset of approximately 600 participants, from whom blood samples were collected at study entry and 14 days post-vaccination.

Antibody responses elicited by MVA-BN-RSV were measured using RSV-specific immunoglobulin (Ig) G enzyme linked immunosorbent assay (ELISA), IgA ELISA and plaque reduction neutralization test for RSV subtype A and subtype B, as described earlier [19]. Serum assays were performed at Bavarian Nordic GmbH, Martinsried, Germany.

### 2.5. Safety assessments

Local and systemic solicited adverse events (AEs) were assessed for 7 days after vaccination, unsolicited AEs were monitored for 28 days post vaccination and beyond if ongoing. Information on SAEs was collected throughout the study. Vital signs (peripheral oxygen saturation, respiratory rate, blood pressure, pulse rate and body temperature) were assessed at on-site study visits. AEs were graded in intensity using the FDA Toxicity Grading Scale for Healthy Adult and Adolescent Volunteers Enrolled in preventive Vaccine Clinical Trials [25].

### 2.6. Statistical methods

The sample size was calculated based on an expected VE of 80% for RSV-associated LRTD with ≥ 3 symptoms, and of 65% for LRTD with ≥ 2 symptoms. Assuming an incidence rate of 0.2% for LRTD with ≥ 3 symptoms, and 0.4% for LRTD with ≥ 2 symptoms, in the placebo group during one RSV season, approximately 20,000 participants vaccinated in the 2 treatment groups allowed the observation of at least 22 cases of LRTD with ≥ 3 symptoms and 53 cases of LRTD with ≥ 2 symptoms, providing at least 90% and 85% power, respectively.

A hierarchical testing strategy was employed. Endpoints were to be tested sequentially against the null hypothesis of VE ≤ 20% at the 2-sided significance level α=0.05, i.e. the lower bound of the 2-sided confidence interval (CI) around the estimate of VE was to be greater than 20% to meet the objective for the respective endpoint.

The full analysis set (FAS) was the primary analysis set for efficacy assessments; it consisted of participants who were randomized to any treatment arm and received the study vaccination. The safety set (SS) was the primary analysis set for safety evaluation; it included participants who received any MVA-BN-RSV vaccine or placebo.

During the study, multiple enrollers were identified through monitoring visits based on similarities in participants details. A systematic verification of all study participants was then performed with support from the vendor who processed participant compensation fees. In total, 826 participants were identified as having been enrolled and vaccinated more than one time in the trial, at different sites located mostly within a 30-mile distance. These participants had received multiple doses of MVA-BN-RSV or placebo (in total 1824 vaccinations). These multiple enrollers were included in the FAS for the primary efficacy analysis with group allocation based on the first treatment received (MVA-BN-RSV or placebo) and with data censored from the time of second vaccination. Multiple enrollers were included in the safety set for safety analysis, with participants having received the MVA-BN-RSV vaccine at least once analyzed in the MVA-BN-RSV group, and those who had only received placebo analyzed in the placebo group.

For the primary analysis, VE against LRTD was defined as the relative reduction in the hazard rate of LRTD associated with RSV in the MVA-BN-RSV vaccinated group compared with that in the placebo group. For each of the 2 primary endpoints, a Cox proportional hazards regression model based on the FAS, stratified by age group, was used as the primary analysis to establish VE against LRTD. The follow-up time for participants who developed LRTD associated with RSV was defined as (first LRTD onset date *minus* vaccination date *plus* 1), and LRTD onset was defined as the date when the first LRTD symptoms were observed, with the sample obtained at the related on-site visit being positive for RSV by PCR. Participants who did not develop any LRTD event were censored at time of dropout or at the end date of the RSV season. The same approach was adopted for the analysis of the key secondary endpoint (ARD associated with RSV).

Post-hoc analyses were performed based on self-reported symptoms in the RiiQ. In the RiiQ, 4 symptoms (coughing, wheezing, shortness of breath and sputum production) constituted the Lower Respiratory Symptoms domain. Based on self-reported symptoms for participants with PCR-confirmed RSV disease, VE against LRTD with at least 2 or 3 self-reported symptoms among the 4 listed, was calculated. In addition, the area under the curve (AUC) of the change from baseline in the RiiQ total score (defined as the mean of the 3 domain scores) and the mean duration of any symptoms were calculated for the 2 treatment groups.

Descriptive summaries per group were provided for local and systemic solicited AEs with any intensity and per intensity grade; unsolicited AEs, including any intensity, grade ≥ 3 and related events classified according to the Medical Dictionary for Regulatory Activities (MedDRA); SAEs and related SAEs.

RSV subtype A and B neutralizing serum activity and RSV-specific IgG, IgA antibody titers at baseline and 14 days post-vaccination were summarized descriptively by treatment group. Geometric mean titers (GMT) and geometric mean fold-increases (GMFI) from baseline were calculated with 95% CIs for all immunogenicity endpoints.

## 3. Results

### 3.1. Study population

A total of 18348 participants were enrolled and vaccinated; multiple enrollers were counted once. Overall, 826 participants were identified as multiple enrollers. These participants were handled appropriately during analyses and consequently did not affect the reported results. A limited number of participants were identified by study sites as having participated in another RSV clinical trial. These participants were excluded from efficacy analyses. The resulting FAS and SS are described in Figure 1.

**Figure 1.**
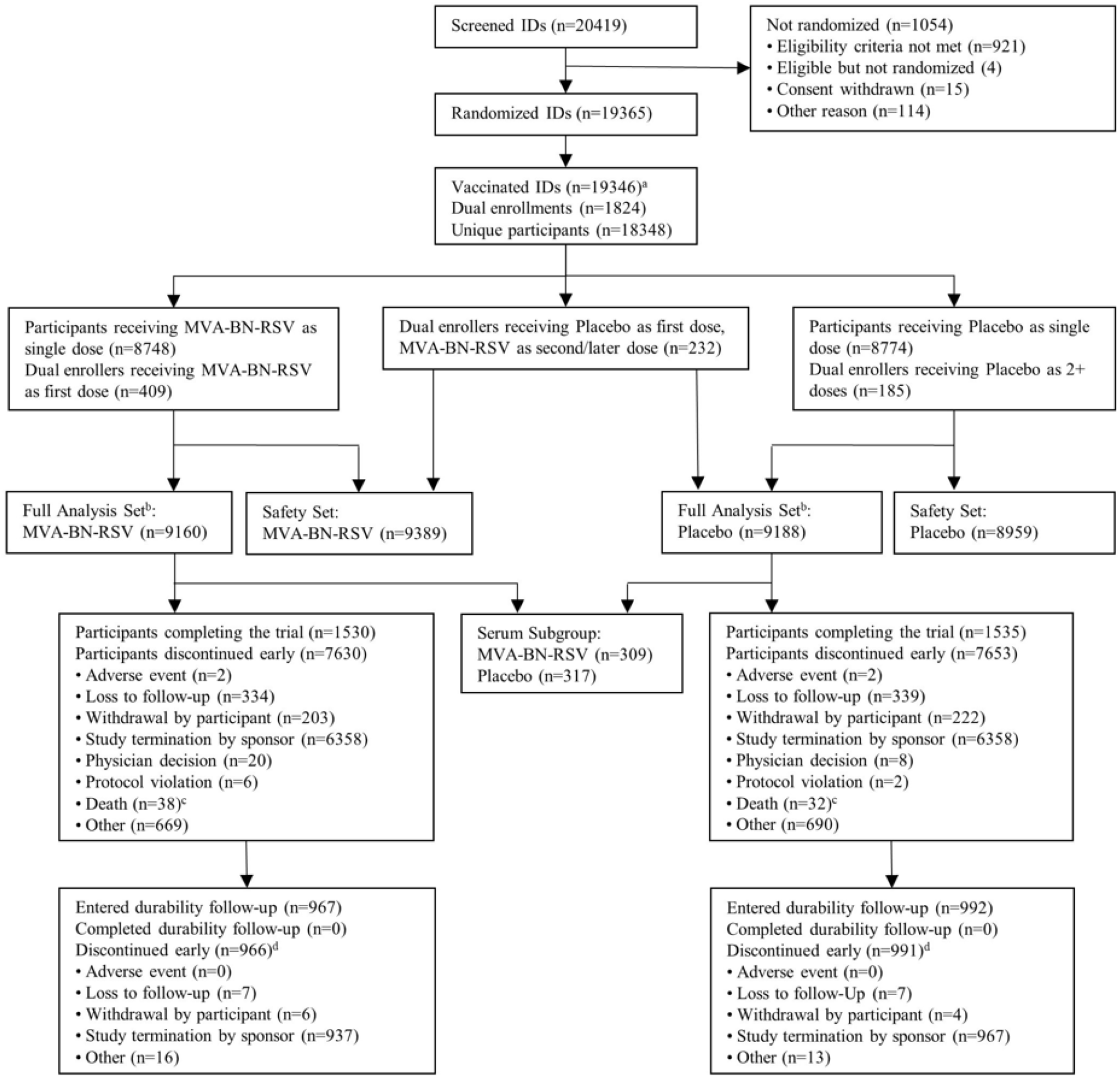
Enrolment and disposition of participants in the analyses. a. Nineteen participants were randomized but not vaccinated; these included 12 additional participant withdrawals, 1 discontinuation of a noncooperative participant, 4 discontinuations after technical difficulties, 1 protocol violation, and 1 discontinuation for ineligibility (participation in another study). b. Treatment allocation based on the (first) randomization; 5 participants received a different treatment than the one they were randomized to (4 participants randomized to MVA-BN-RSV received placebo and 1 participant randomized to placebo received MVA-BN-RSV instead). c. There were 5 additional deaths reported in the study: 2 deaths were reported after participant withdrawal and 1 after study completion. The other 2 participants were dual/multiple enrollers and were reported as withdrawn or discontinued from their first enrolment; their deaths were reported with a later enrolment. d. One participant in each treatment group confirmed consent for durability follow-up but did not have any activity and therefore were not counted as having discontinued durability follow-up

The demographic characteristics of participants were similar in the 2 groups (Table 1). Their mean age was 70.4 years, more than 4000 (22.5%) were ≥ 75 years of age, 66.5% had underlying chronic lung or cardiac diseases putting them at higher risk of severe RSV disease and 10.7% were frail. Overall, 13.0% of study participants were Black or African American and 28.7% were Hispanic or Latino.

**Table 1.**
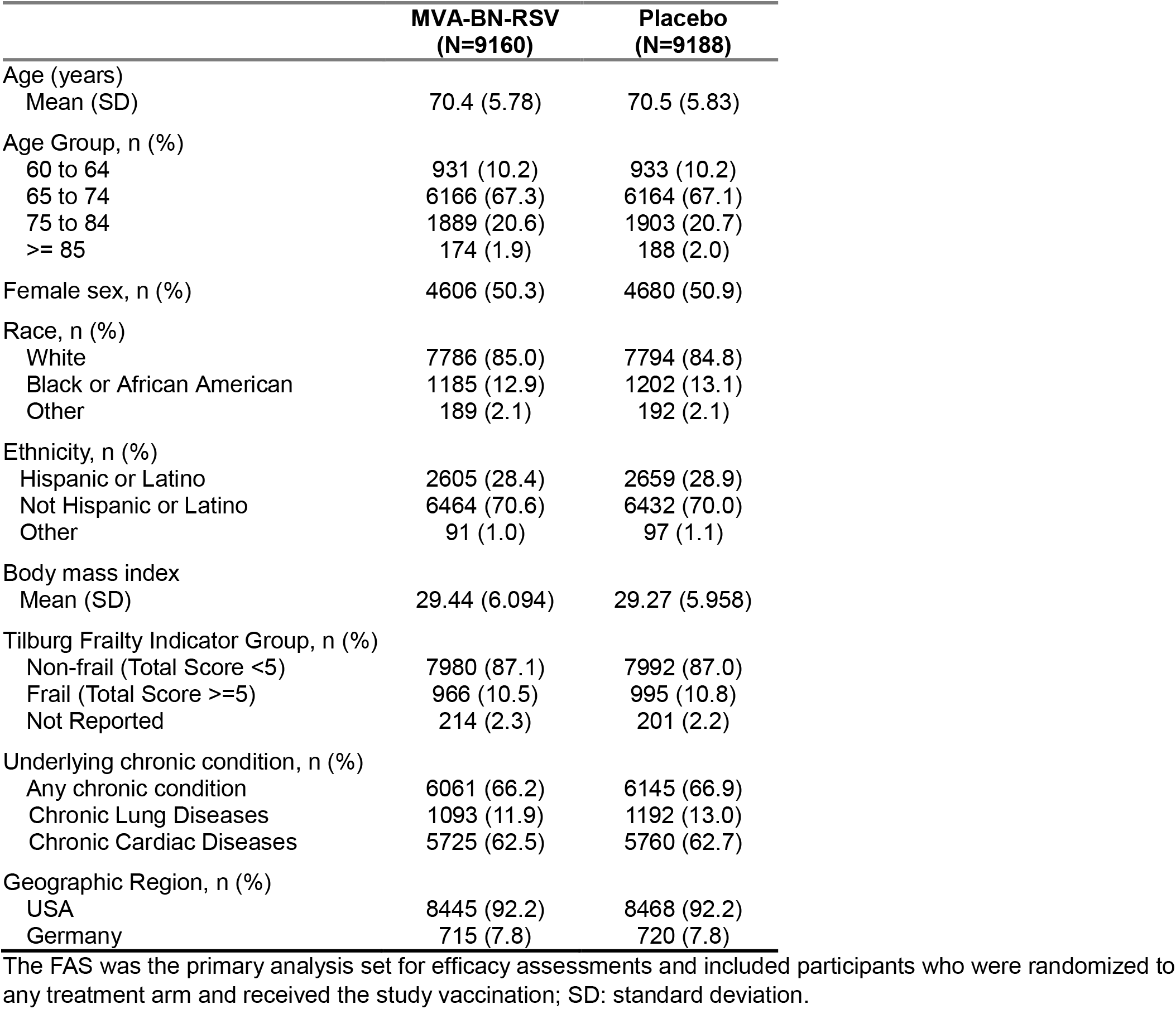
Demographics and baseline characteristics of participants (FAS)

### 3.2. Efficacy

The assumption used for sample size calculation of a 0.2% incidence for LRTD with ≥ 3 symptoms during one RSV season was confirmed since 21 cases were reported for 9188 placebo recipients (0.23%) during the RSV season. The cut-off date for the analysis of efficacy was 6 March 2023, at which time, the RSV season was considered over based on country-specific surveillance data [26, 27]. The number of LRTD events accrued by that time was above the number required by the sample size estimation.

There were 12 reports of RSV-associated LRTD with ≥ 3 symptoms in the MVA-BN-RSV group and 21 in the placebo group, giving a VE of 42.9% (95% CI: -16.1; 71.9) (Figure 2A). For this primary endpoint, the study failed to meet the success criterion of a lower bound of the 95% CI greater than 20%. There were 25 reports of RSV-associated LRTD with ≥ 2 symptoms in the MVA-BN-RSV group and 61 in the placebo group, for a VE of 59.0% (95% CI: 34.7; 74.3) (Figure 2B). For RSV-associated ARD, VE was 48.8% (95% CI: 25.8; 64.7) with 42 reports in the MVA-BN-RSV group and 82 in the placebo group (Figure 3C). For LRTD with ≥ 2 symptoms and ARD, the lower bound of the 95% CI was above 20%. Per the planned hierarchical testing strategy, no formal conclusion can be drawn for these 2 endpoints, although positive efficacy was observed.

**Figure 2.**
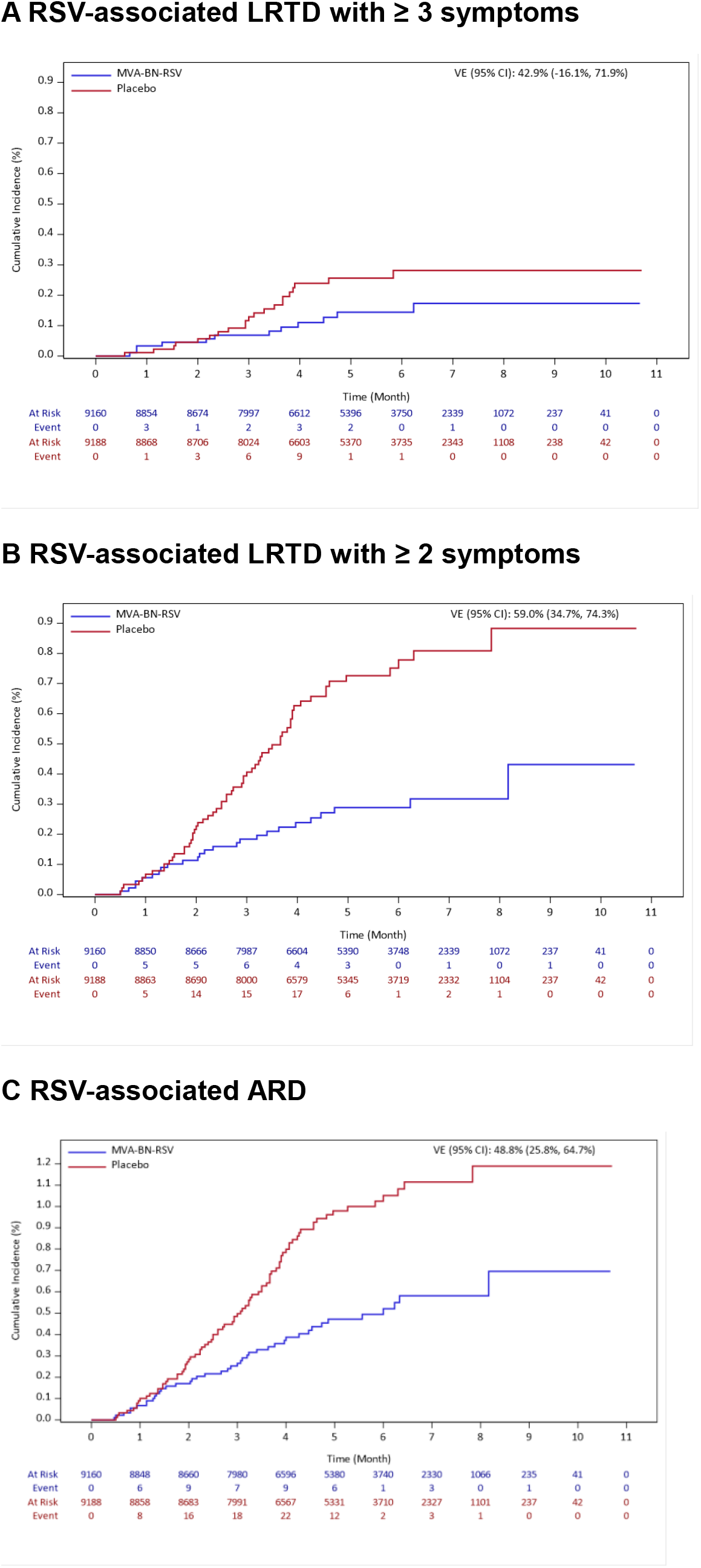
**Cumulative incidence of RSV-associated LRTD and RSV-related ARD (FAS)** This Figure presents the cumulative incidence of RSV–associated lower respiratory tract disease with at least 3 symptoms (Panel A), with at least 2 symptoms (Panel B) and RSV-associated acute respiratory disease (Panel C). The analysis included PCR-confirmed RSV-associated LRTD or ARD with onset at least 14 days after vaccination (full analysis set). The full analysis set was the primary analysis set for efficacy assessments; it consisted of participants who were randomized to any treatment arm and received the study vaccination.

**Figure 3.**
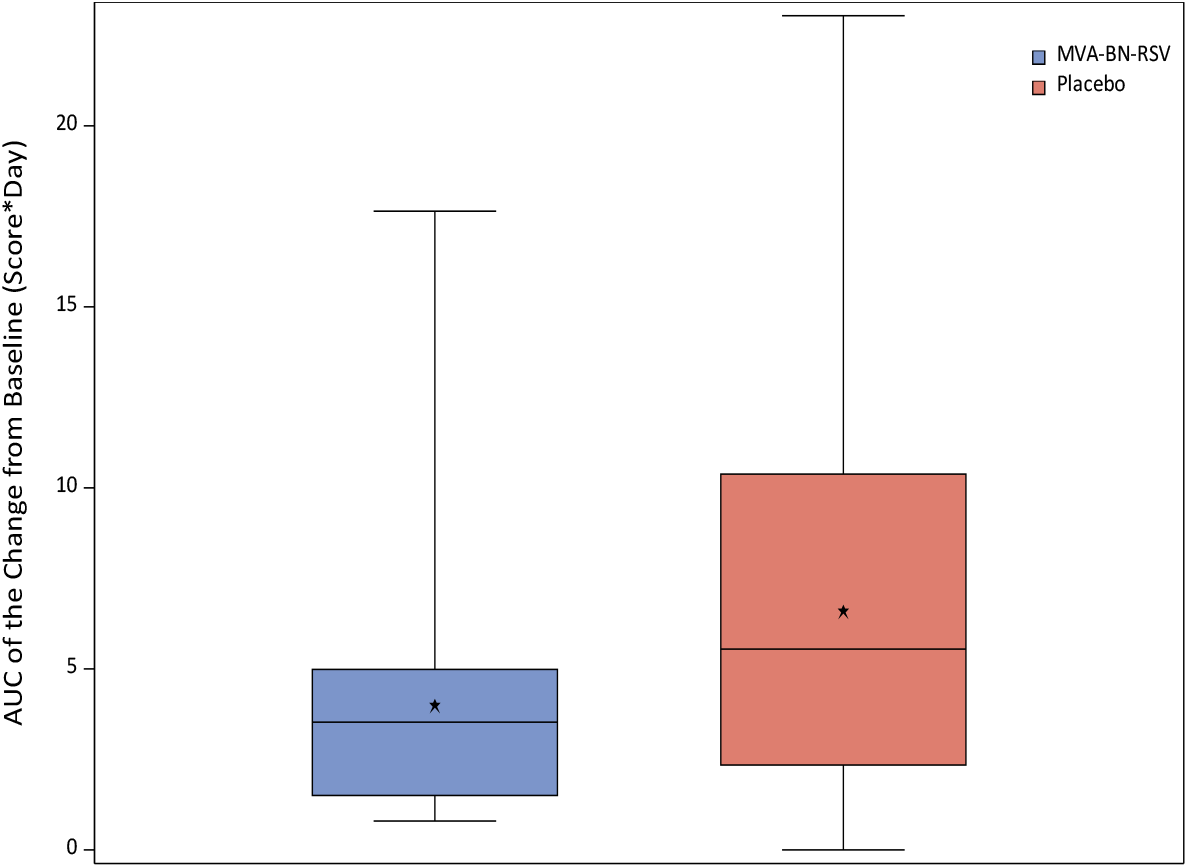
**Severity of RSV-associated LRTD symptoms based on RiiQ scores** This graph shows the total respiratory and systemic symptom severity among participants with RT-PCR–confirmed RSV-associated acute respiratory disease as measured with the Respiratory Infection Intensity and Impact Questionnaire (RiiQ); higher area-under-the-curve (AUC) values indicate greater symptom severity. The top and bottom of the box indicate the third and first quartiles, the whiskers indicate the range (minimum, maximum), the horizontal line inside the box indicates the median; the star indicates the mean value.

There were no hospitalizations due to RSV-associated disease during the study. There were 2 cases of RSV-associated LRTD that met the definition of severe disease, both in the placebo group. One had a respiratory rate > 25 breaths/Min and one had imaging evidence of new onset of pneumonia.

The exploratory analysis of vaccine efficacy based on the number and intensity of symptoms self-reported by participants in the RiiQ confirmed a moderate efficacy against RSV-associated LRTD, which ranged from 58.1% (95% CI: 36.3; 72.5) to 77.8% (95% CI: 56.0; 88.8), depending on the number and severity of reported symptoms (Supplementary Table 1). The analysis of the severity of symptoms of RSV-associated disease based on the RiiQ showed that RSV-associated LRTD was milder in RSV vaccine recipients who experienced breakthrough disease (Figure 3). The median duration of symptoms was also lower in the RSV-MVA-BN group (11.5 days) than in the placebo group (15 days).

Subgroup analyses were consistent with the findings of the primary efficacy analysis (data on file), except for the analysis per sex, which showed higher VE estimates in females compared to males, for the primary and key secondary RSV disease endpoints (Supplementary Table 2). However, while VE was substantially higher in female participants compared to male participants in the subset with exactly 1 symptom and the subset with 3 or more symptoms, it was almost the same in the subset with exactly 2 symptoms: 69.7% (95% CI: 24.5; 87.8) in females *versus* 65.3% (95% CI: 18.0; 85.3) in males (Supplementary Table 3).

### 3.3. Immunogenicity

MVA-BN-RSV induced a humoral immune response in terms of neutralizing antibodies against the 2 RSV subtypes, and of RSV-specific IgG and IgA (Table 2). Fourteen days after RSV-MVA-BN vaccination, GMFIs were 1.7 for RSV A and RSV B neutralizing antibody titers, and 2.9 and 4.3 for IgG and IgA antibody titers, respectively. The analysis of immune responses per sex (Supplementary Table 4) showed slightly higher baseline titers and slightly lower fold increases in males for all immunogenicity measurements.

**Table 2.**
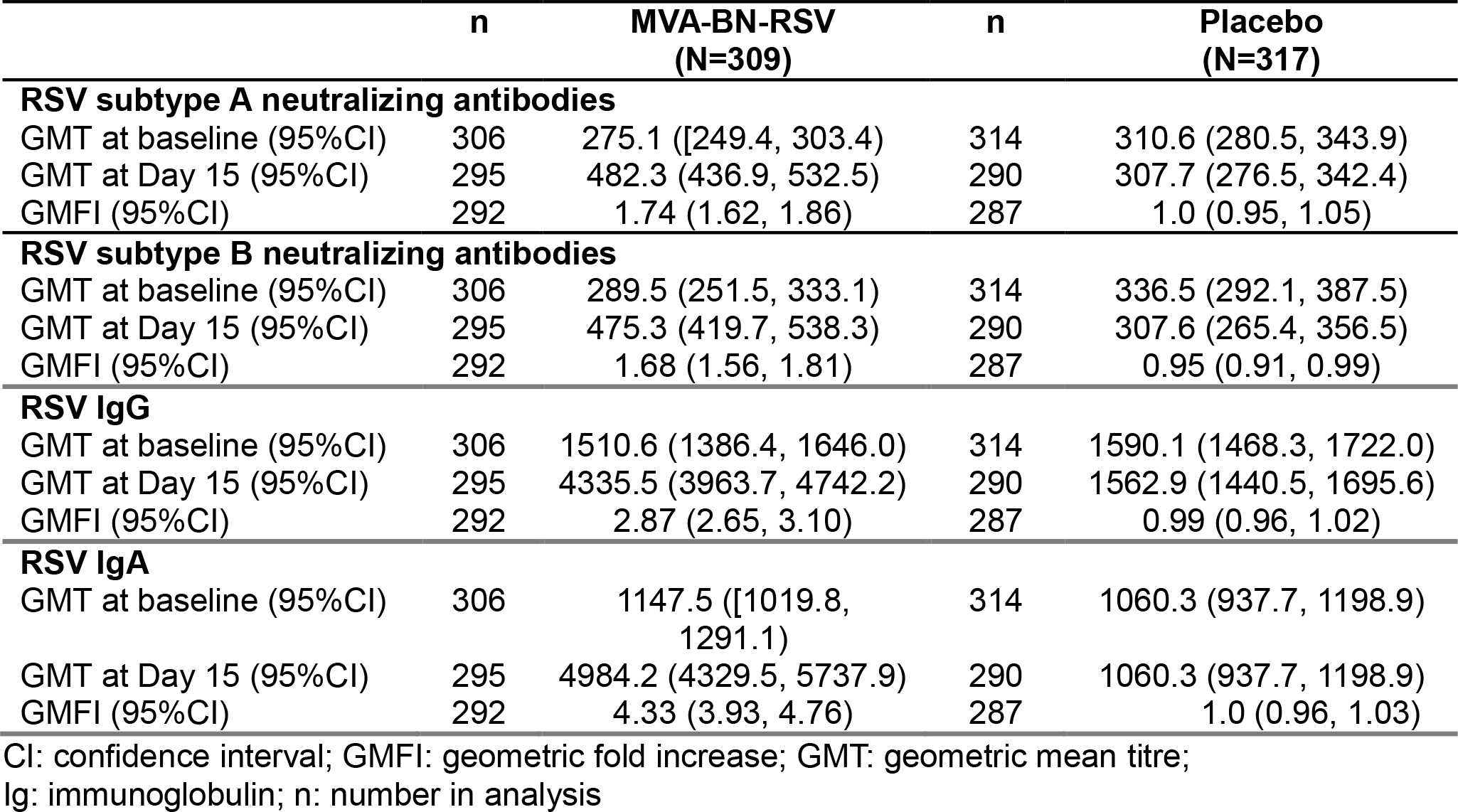
Humoral immune response at baseline and 14 days post-vaccination (serum subset)

### 3.4. Reactogenicity and safety

The most frequent solicited AEs, reported by more than 30% of vaccinees, were pain at the injection site, myalgia, fatigue, and headache (Figure 4). Grade 3 solicited local AEs were reported by 4.6% of MVA-BN-RSV vaccinees, and grade ≥ 3 systemic AEs by 4.5% of vaccine recipients. A similar proportion of participants (6.5%) in the 2 groups reported unsolicited AEs during the 28 days following vaccination (Table 3); and during that same period, 1.1% participants in the MVA-BN-RSV group compared to 0.6% in the placebo group reported related unsolicited AEs. The reported unsolicited AEs were generally mild (Grade 1) and self-limited. Grade ≥ 3 unsolicited AEs assessed as related to study intervention were consistent with the solicited AEs reported following vaccination by participants. Vital sign monitoring did not detect any physical findings of concern.

**Figure 4a.**
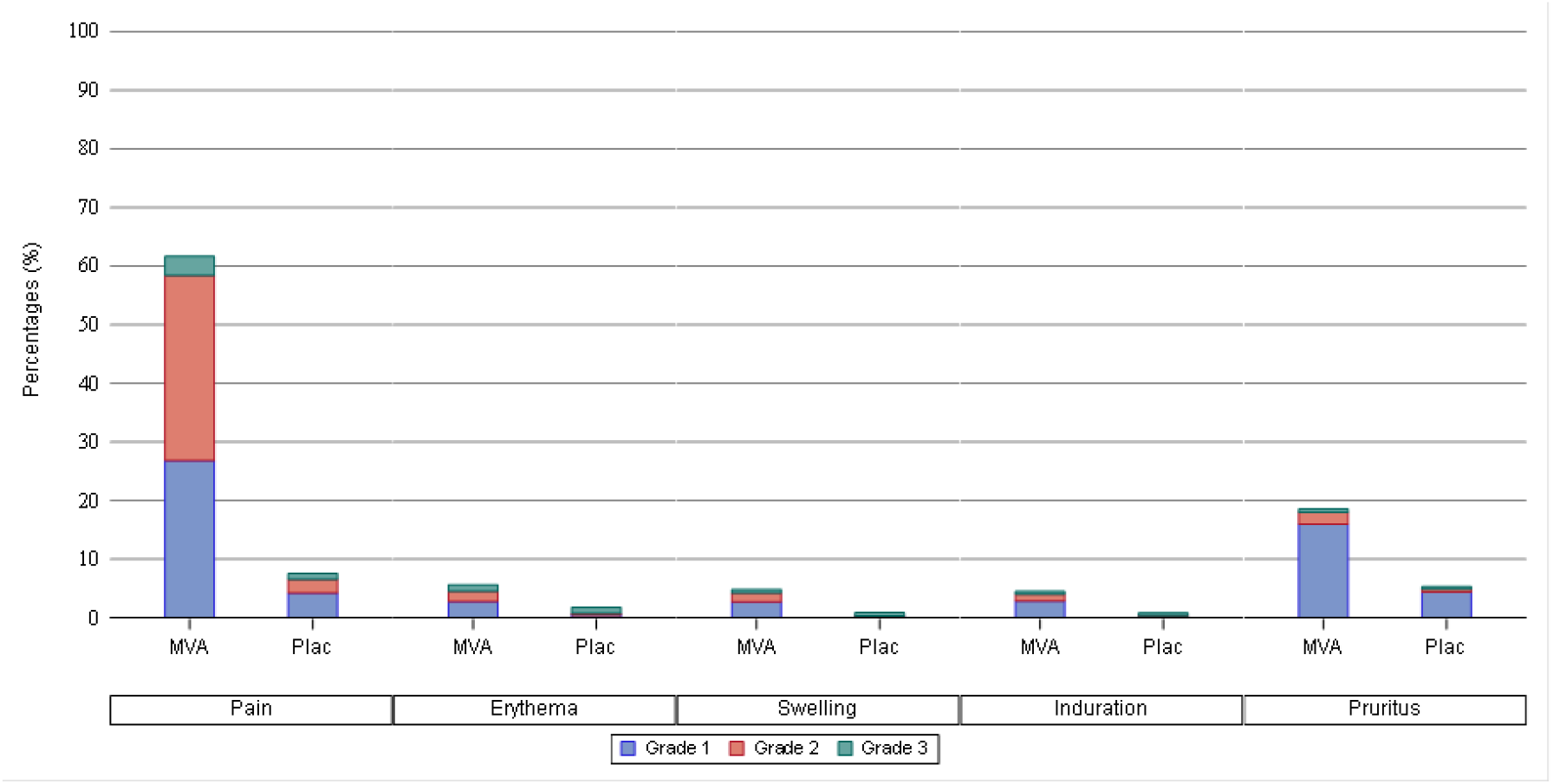
Frequency of Solicited Local Adverse Events after vaccination (SS)

**Figure 4b.**
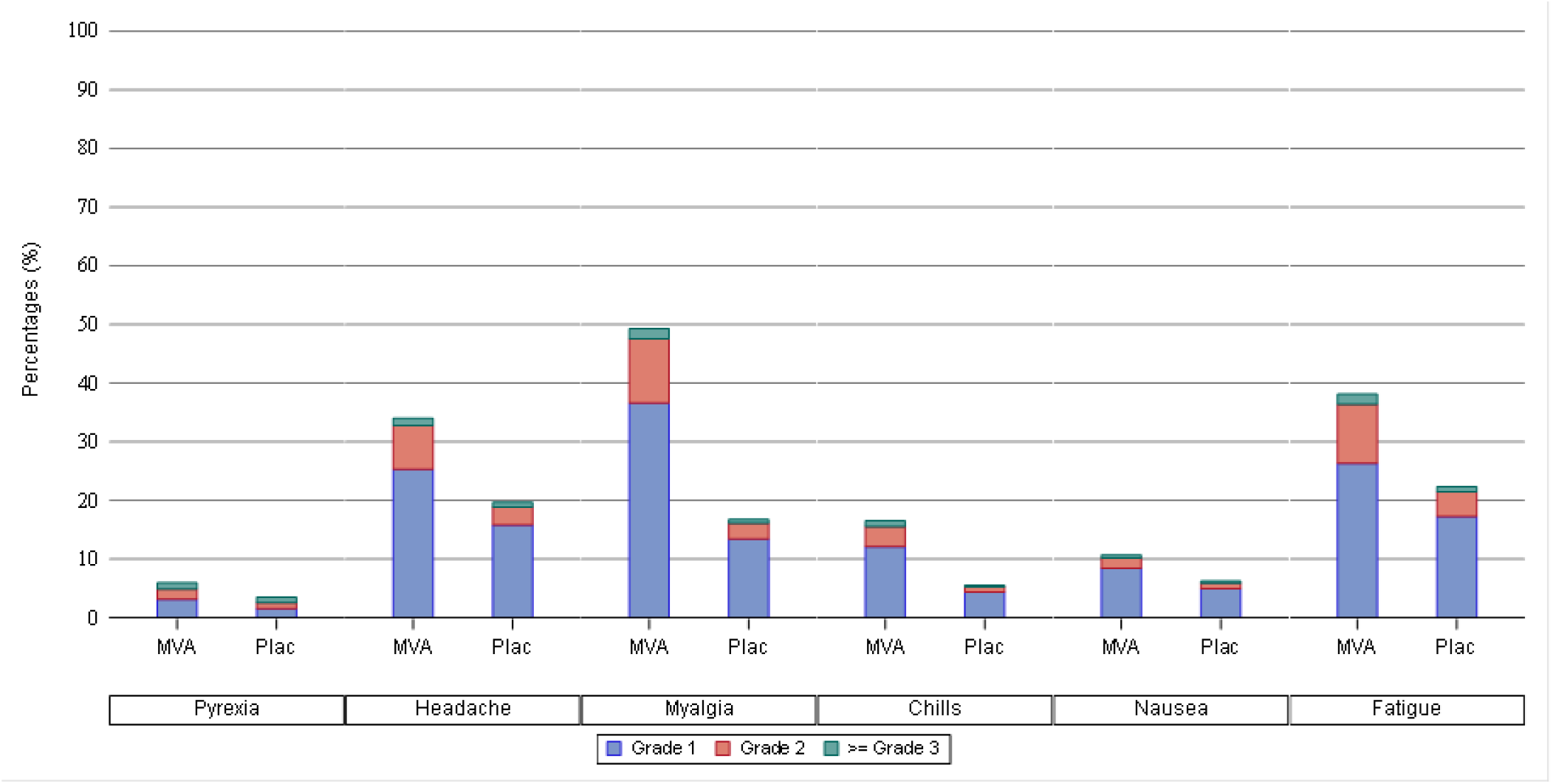
Frequency of Solicited Systemic Adverse Events after vaccination (SS) The safety set was the primary analysis set for safety evaluation; it included participants who received any MVA-BN-RSV vaccine or placebo. MVA: MVA-BN-RSV group; Plac: placebo group; Percentage: percentage of participants reporting an AE. In Figure 4a, solicited local AEs were pain, erythema, swelling, induration and pruritus reported at the injection site during the 7 days following vaccination. In Figure 4b, solicited systemic AEs were pyrexia, headache, myalgia, chills, nausea and fatigue reported during the 7 days following vaccination.

**Table 3.** Frequency of Unsolicited Adverse Events and SAEs after vaccination (SS)

|  | MVA-BN-RSV<br>(N=9389) | Placebo<br>(N=8959) |
| --- | --- | --- |
| **Unsolicited Adverse Events** (within 28 days after vaccination) |  |  |
| All, n (%) | 607 (6.5) | 581 (6.5) |
| Grade ≥3, n (%) | 71 (0.8) | 45 (0.5) |
| Related, n (%) | 103 (1.1) | 50 (0.6) |
| Grade≥3 & Related, n (%) | 11 (0.1) | 2 (<0.05) |
| **Serious Adverse Events** (during the entire study period) |  |  |
| All, n (%) | 517 (5.5) | 422 (4.7) |
| Related, n (%) | 7 (0.1) | 5 (0.1) |
| Fatal, n (%) | 41 (0.4) | 34 (0.4) |

The safety set was the primary analysis set for safety evaluation; it included participants who received any MVA-BN-RSV vaccine or placebo. Related: thought to be related to study vaccination or placebo by the investigator.

SAEs collected during the entire study period were reported with similar frequencies in the 2 study groups: 5.5% and 4.7% in the MVA-BN-RSV group and in placebo group, respectively. SAEs considered to be related to the study vaccination (possibly, probably, or definitely related to study vaccination) occurred in 7 participants, (0.1%, 10 events) in the MVA-BN-RSV group, and in 5 participants (0.1%, 5 events) in the placebo group. In both groups, 1 event each of atrial fibrillation and pneumonia was reported. In the MVA-BN-RSV group, there were also 2 events of cerebrovascular accident, 2 events of syncope, 1 event of an altered state of consciousness, 1 event of pyrexia, and 1 participant presented the events of injection site erythema and pruritus. In the placebo group, in addition, 1 event each of colitis, wheezing, and thrombosis occurred. No fatal cases were considered to be related to the study vaccination. A total of 75 participants died during the study: 41 (0.4%) in the MVA-BN-RSV group and 34 (0.4%) in the placebo group. The most frequently reported fatal SAE by System Organ Class was cardiac disorder in the 2 study groups. The causes of death reported among study participants were representative of the most common causes of death in the elderly adult population and, considering that these fatal events were more likely attributable to the participant’s underlying risk factors (e.g. diabetes mellitus type 2, hypertension), none of the fatal events was considered by the study investigator to be related to study vaccination. The safety information was continuously presented to the Data Monitoring Committee and no concern was raised after review and assessment.

## 4. Discussion

This study was undertaken to assess the efficacy of the MVA-BN-RSV vaccine against RSV-associated ARD and LRTD in adults ≥ 60 years of age.

The study population was reflective of the general population of older adults targeted for RSV vaccination, as more than 4000 participants were ≥ 75 years of age, a majority had underlying cardiac or respiratory conditions and 10.7% were frail. The study population reasonably reflected racial and ethnic diversity in the study locations and was well balanced for sex and other demographic characteristics across the two groups.

VE against LRTD with at least 3 symptoms was 42.9% (95% CI: -16.1; 71.9) and the predefined success criterion of a CI lower bound greater than 20% was not met. Given this, per the hierarchical testing strategy adopted, no formal conclusion on efficacy relating to the endpoints tested next in the sequence can be made. The results of the analyses for VE against RSV-associated LRTD with at least 2 symptoms and against ARD showed estimates of 59.0% and 48.8% respectively with lower bounds above 20%, and are consistent with a positive, although suboptimal, protection against RSV disease. The analysis of the patient-reported outcomes through the RiiQ also confirmed the presence of efficacy against RSV-associated disease, with moderate estimates that were in line with those reported in the primary analysis for LRTD with at least 2 symptoms. The fact that the patient-reported severity and duration of symptoms were lower in breakthrough cases in the vaccine group than in the placebo group also supports the fact that the vaccine conferred some protection against RSV-associated disease, although not to the expected level.

Subgroup analyses of VE for each of the disease endpoints per age, sex, race, underlying chronic conditions, and geographic regions were performed (data on file). These analyses were generally consistent with the primary analyses, except for the per sex analysis, which showed higher VE estimates in females. The vaccine immunogenicity measured in this study was in line with Phase 1 and 2 assessments, showing comparable fold-increases and post-vaccination titers in the two sexes [18, 19]. In this study, the analysis of immunogenicity per sex did not reveal major differences between males and females, although slightly higher baseline levels, and thus slightly lower fold increases, were seen in males. Also, in earlier studies [18, 19], cellular responses analyzed per sex showed similar values and fold-increases in males and females (data on file). Other efficacy studies of RSV vaccines did not report a higher efficacy in females [10, 28, 29], and the difference in VE across sexes observed in this trial was not expected. In view of the similar immunogenicity and baseline characteristics in males and females (data on file), we have not found an explanation, nor a plausible physiological mechanism for a differential efficacy per sex for this vaccine. Generally, females, including elderly women, usually mount a higher immune response to vaccines than males, and vaccine efficacy in females tends to be higher regardless of age or vaccine [30]. A study evaluating sex effect in influenza vaccine effectiveness showed a 19% (48% for females and 29% for males, p = 0.03) difference in adults ≥50 years [31], although vaccine efficacy is generally not considered different between females and males for available influenza vaccines. A review article investigating sex differences in respiratory viral pathogenesis of disease revealed males to be more susceptible than females to severe outcomes from respiratory viral infections at younger and older ages in general, but no sex specific data was found for RSV-associated disease in older adults [32].

Given that VE was substantially higher in females than in males for the subset with ≥ 3 symptoms, it may be possible that MVA-BN-RSV, a unique vaccine that expresses 5 RSV proteins, proved more effective in females in preventing more severe disease, which would be corroborated by the fact that, for LRTD with exactly 2 symptoms, VE was similar (65-70%) in the two sexes. However, VE was also higher in females compared to males for the mild disease subset (i.e. with ≤ 1 symptom), which does not validate the view that the sex difference would be limited to more severe disease. Due to the small number of cases in subgroups and the lack of identified biological plausibility, we cannot preclude the influence of other random factors.

In our study, only 2 cases of RSV-associated disease with complications or of high severity were reported, both in the placebo group; due to the low number of cases, this does not inform on the level of protection that MVA-BN-RSV could afford against severe RSV disease.

Before initiating vaccine efficacy evaluation in this Phase 3 study, the ability of MVA- BN-RSV to prevent RSV infection and colonization was assessed in an RSV challenge trial, where MVA-BN-RSV vaccination was effective at reducing the RSV viral load, the number of RSV infections and symptoms after challenge [20], and vaccine efficacy against RSV- confirmed, symptomatic infection was 79.3% to 88.5%. These values were not reproduced in the efficacy study reported here. The RSV controlled human challenge trials have inherent limitations due to the use of highly attenuated RSV strains causing mild to moderate upper respiratory tract disease; such studies can reveal the protective potential of a vaccine, though their results cannot be directly reflective of larger vaccine efficacy studies or real-world analysis [33]. The results of the MVA-BN-RSV challenge trial are nevertheless coherent with the efficacy estimated against ARD and milder LRTD with at least 2 symptoms, but do not align with the efficacy seen against more severe LRTD.

The adopted vaccine approach proved unable to afford the pursued high level of protection despite the strong and diverse T cell response elicited, which, given the recognized importance of T cell responses for protection against RSV symptomatic infection [34], had given confidence in the multi antigen approach. This broad T cell response was not capable of counterbalancing a lower neutralizing antibody response, confirming the importance of a high neutralizing antibody response and preF focus for optimal protection against RSV. Indeed, the humoral immune response elicited by MVA-BN-RSV was lower than that elicited by preF- focused vaccines. Although assays are different and values not directly comparable, the fold-increases from baseline (1.7 for neutralizing antibodies and 2.9 for IgG) were lower than those described for preF-based vaccines, which ranged 8-12 for neutralizing antibodies [8, 14, 35] and 8-13 for IgG [8, 14]. The efficacy observed for MVA-BN-RSV, was suboptimal but was higher than that reported with two postF-based vaccines. These showed no efficacy in Phase 3: one displayed a VE of -7.1% (90% CI: -106.9; 44.3) against RSV-associated acute respiratory infection [36], and the other VEs of -7.9% (unspecified% CI: -8; 37) against RSV-associated moderate-severe LRTD, and 12.6% (unspecified% CI: -14; 33) against ARD [37]. This is in contrast with the preF-focused vaccines, which all showed high efficacy in the initial first RSV season evaluation regardless of the platform [8, 9, 10, 14]. MVA-BN-RSV, which induces a mixed pre- and post-fusion expression of the RSV F protein, therefore appears to confer an intermediate efficacy between that afforded by the postF-based and the preF-based vaccines.

From an operational point of view, although the presence of participants who had enrolled more than once and had received multiple vaccinations had no bearing on the analysis, thanks to early detection and appropriate handling, the issue of multiple enrollers in clinical studies is of concern, as this has potential to impair results, and should be a point of attention in the conduct of trials.

The evaluation of MVA-BN-RSV in more than 9000 older individuals in this study confirmed the mild to moderate reactogenicity profile of the vaccine. A similar number of adverse events were observed among the 2 study groups and no safety concern was identified by the Data Monitoring Committee. These data are in line with those obtained with the MVA-BN poxvirus vaccine Jynneos® [38]. In all completed clinical trials, vaccinations with MVA-BN or MVA-BN-based vaccines displayed a good safety profile and were well tolerated in all the populations of healthy, elderly or immunocompromised individuals assessed. A recent meta-analysis encompassing 8 clinical studies with MVA-BN, of which 6 were placebo-controlled, showed that, following a single dose of MVA-BN, no difference with placebo was demonstrated for the outcomes of SAEs, AEs of special interest or AEs requiring discontinuation [39].

## 5. Conclusions

The MVA-BN-RSV vaccine demonstrated suboptimal efficacy against RSV-associated LRTD, in this double-blind placebo-controlled randomized trial. Although efficacy against the first primary endpoint was not met, the vaccine displayed some, moderate protective efficacy against RSV-associated LRTD and ARD. The immunogenicity of the vaccine was also established, confirming that MVA-BN can be used as a vector platform, to build safe and effective vaccines based on adequate constructs. This large trial also confirmed the safety of the MVA-BN platform as a vector, including in an older population with co-morbidities.

All authors attest they meet the ICMJE criteria for authorship.

## Data Availability

All data produced in the present study are available upon request to the authors

## Acknowledgments

We thank the participants of the clinical study, the clinical site investigators and their staff and the ICON clinical research team.

## Financial support

This work was supported by Bavarian Nordic A/S.

---

**Supplementary Table 1.** Occurrence and Vaccine Efficacy of LRTD based on RiiQ Responses

| Number of LRTD Symptoms in RiiQ | MVA-BN-RSV<br>N=9160<br>n events | Placebo<br>N=9188<br>n events | Vaccine Efficacy<br>% (95% CI) |
| --- | --- | --- | --- |
| ≥3 mild symptoms | 18 | 51 | 64.8 (39.7; 79.4) |
| ≥2 mild symptoms | 31 | 74 | 58.1 (36.3; 72.5) |
| ≥3 moderate symptoms | 6 | 19 | 68.5 (21.1; 87.4) |
| ≥2 moderate symptoms | 10 | 45 | 77.8 (56.0; 88.8) |

LRTD: lower respiratory tract disease; RiiQ: Respiratory Intensity and Impact Questionnaire; CI: confidence interval

**Supplementary Table 2.** Vaccine Efficacy against RSV-associated LRTD and ARD – Analysis per Sex (Full Analysis Set)

|  | N | MVA-BN-RSV |  | N | Placebo |  | Hazard Ratio<br>(95% CI) | Vaccine Efficacy<br>% (95% CI) |
| --- | --- | --- | --- | --- | --- | --- | --- | --- |
|  |  | n events (%) | Censor, n (%) |  | n events (%) | Censor, n (%) |  |  |
| **LRTD ≥ 3 symptoms** |  |  |  |  |  |  |  |  |
| **Male** | 4554 | 10 (0.2) | 4544 (99.8) | 4508 | 7 (0.2) | 4501 (99.8) | 1.415<br>(0.539, 3.718) | -41.5<br>(-271.8, 46.1) |
| **Female** | 4606 | 2 (<0.05) | 4604<br>(100.0) | 4680 | 14 (0.3) | 4666 (99.7) | 0.145<br>(0.033, 0.636) | 85.5<br>(36.4, 96.7) |
| **LRTD ≥ 2 symptoms** |  |  |  |  |  |  |  |  |
| **Male** | 4554 | 17 (0.4) | 4537 (99.6) | 4508 | 27 (0.6) | 4481 (99.4) | 0.623<br>(0.340, 1.144) | 37.7<br>(-14.4, 66.0) |
| **Female** | 4606 | 8 (0.2) | 4598 (99.8) | 4680 | 34 (0.7) | 4646 (99.3) | 0.238<br>(0.110, 0.514) | 76.2<br>(48.6, 89.0) |
| **ARD** |  |  |  |  |  |  |  |  |
| **Male** | 4554 | 28 (0.6) | 4526 (99.4) | 4508 | 33 (0.7) | 4475 (99.3) | 0.841<br>(0.508, 1.391) | 15.9<br>(-39.1, 49.2) |
| **Female** | 4606 | 14 (0.3) | 4592 (99.7) | 4680 | 49 (1.0) | 4631 (99.0) | 0.288<br>(0.159, 0.522) | 71.2<br>(47.8, 84.1) |

LRTD: lower respiratory tract disease; ARD: Acute respiratory disease; CI: confidence interval

**Supplementary Table 3.**
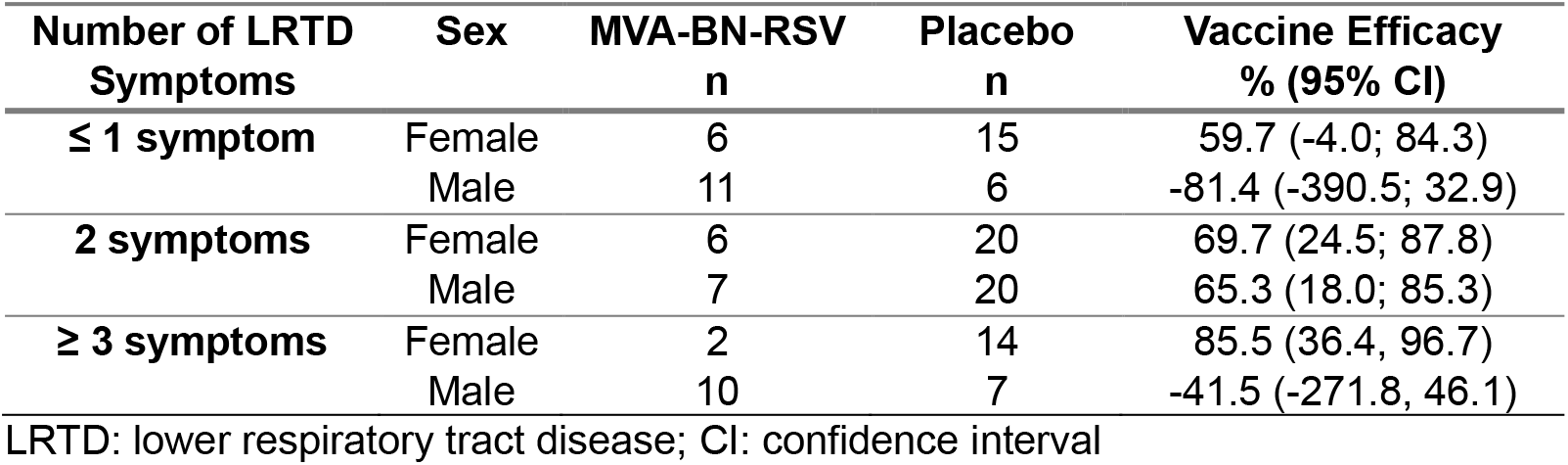
Vaccine efficacy against RSV-associated LRTD – post hoc analysis per sex, per exact number of symptoms (FAS)

**Supplementary Table 4.** Humoral immune response at baseline and 14 days post-vaccination (serum subset) - per sex analysis

| Sex | Parameter | Group | Visit | N | GMT | GMT<br>(95% CI) | GMFI | GMFI<br>(95% CI) |
| --- | --- | --- | --- | --- | --- | --- | --- | --- |
| Female | RSV IgA (GMT) | MVA-BN-RSV | Baseline | 162 | 960.2 | (825.4; 1117.1) | . |  |
|  |  |  | Day 15 | 157 | 4347.1 | (3609.0; 5236.1) | 4.51 | (3.94; 5.16) |
|  |  | Placebo | Baseline | 147 | 1015.2 | (851.0; 1211.2) | . |  |
|  |  |  | Day 15 | 142 | 992.1 | (832.6; 1182.1) | 1.01 | (0.96; 1.06) |
|  | RSV IgG (GMT) | MVA-BN-RSV | Baseline | 162 | 1370.8 | (1211.3; 15510.3) | . |  |
|  |  |  | Day 15 | 157 | 4069.3 | (3560.3; 4651.1) | 2.95 | (2.64; 3.29) |
|  |  | Placebo | Baseline | 147 | 1448.7 | (1288.3; 1629.0) | . |  |
|  |  |  | Day 15 | 142 | 1407.6 | (1253.1; 1581.1) | 0.98 | (0.95; 1.03) |
|  | RSV Type A<br>Neutralizing GMT | MVA-BN-RSV | Baseline | 162 | 236.4 | (204.6; 273.0) | . |  |
|  |  |  | Day 15 | 157 | 423.9 | (369.9; 485.8) | 1.75 | (1.59; 1.93) |
|  |  | Placebo | Baseline | 147 | 283.5 | (248.6; 323.4) | . |  |
|  |  |  | Day 15 | 142 | 271.3 | (234.1; 314.4) | 0.99 | (0.93; 1.05) |
|  | RSV Type B<br>Neutralizing GMT | MVA-BN-RSV | Baseline | 162 | 278.3 | (229.5; 337.5) | . |  |
|  |  |  | Day 15 | 157 | 429.6 | (360.3; 512.2) | 1.58 | (1.44; 1.73) |
|  |  | Placebo | Baseline | 147 | 267.0 | (219.8; 324.4) | . |  |
|  |  |  | Day 15 | 142 | 255.8 | (209.5; 312.4) | 0.96 | (0.90; 1.03) |
| Male | RSV IgA (GMT) | MVA-BN-RSV | Baseline | 144 | 1402.0 | (1170.7; 1679.1) | . |  |
|  |  |  | Day 15 | 138 | 5823.3 | (4703.6; 7209.5) | 4.13 | (3.60; 4.74) |
|  |  | Placebo | Baseline | 167 | 1155.8 | (980.9; 1334.6) | . |  |
|  |  |  | Day 15 | 148 | 1130.1 | (949.9; 1344.6) | 0.99 | (0.94; 1.04) |
|  | RSV IgG (GMT) | MVA-BN-RSV | Baseline | 144 | 1685.1 | (1499.1; 1894.1) | . |  |
|  |  |  | Day 15 | 138 | 4659.6 | (4143.6; 5239.8) | 2.78 | (2.49; 3.10) |
|  |  | Placebo | Baseline | 167 | 1726.0 | (1549.1; 1923.2) | . |  |
|  |  |  | Day 15 | 148 | 1727.9 | (1542.9; 1935.1) | 1.00 | (0.96; 1.04) |
|  | RSV Type A<br>Neutralizing GMT | MVA-BN-RSV | Baseline | 144 | 326.3 | (287.5; 370.2) | . |  |
|  |  |  | Day 15 | 138 | 558.6 | (484.7; 643.7) | 1.71 | (1.56; 1.88) |
|  |  | Placebo | Baseline | 167 | 336.5 | (288.7; 392.2) | . |  |
|  |  |  | Day 15 | 148 | 347.1 | (297.6; 404.9) | 1.01 | (0.93; 1.09) |
|  | RSV Type B<br>Neutralizing GMT | MVA-BN-RSV | Baseline | 144 | 302.6 | (245.9; 372.3) | . |  |
|  |  |  | Day 15 | 138 | 533.3 | (447.5; 635.7) | 1.80 | (1.61; 2.02) |
|  |  | Placebo | Baseline | 167 | 412.4 | (337.7; 503.5) | . |  |
|  |  |  | Day 15 | 148 | 367.0 | (296.1; 455.0) | 0.94 | (0.90; 1.00) |

GMT: geometric mean titer; GMFI: Geometric mean fold-increase; CI: confidence interval

## Notes

### Competing Interest Statement

Authors are employees at Bavarian Nordic, the sponsor of the clinical trial except Prof Welte who was the principal investigator in Germany

### Clinical Trial

EudraCT 2021-004923-34, NCT05238025

### Funding Statement

The study was funded by Bavarian-Nordic

### Author Declarations

Ethics committee/IRB gave ethical approval for this work, trial was approved by: IRB: WCG IRB, 1019 39th Avenue SE, Suite10 Puyllup, WA98374-2115 (IRB tracking no 20220860) IRB: Ethikkommission Medizinische Hochschule Hannover, Carl-Neuberg-Str 1, 30625 Hannover, Germany (Nr. 10289 AMG M 2022)

## References

1. Piralla A, Chen Z, Zaraket H. An update on respiratory syncytial virus. MC Infectious Diseases (2023) 23:734.

2. Shi T, Denouel A, Tietjen AK, Campbell I, Moran E, Li X, Campbell H, Demont, Nyawanda BO. Chu HY et al. Global Disease Burden Estimates of Respiratory Syncytial Virus–Associated Acute Respiratory Infection in Older Adults in 2015: A Systematic Review and Meta-Analysist al. The Journal of Infectious Disease, 2020;222(S7):S577–83

3. Savic M, Penders Y, Shi T, Branche A, Pirçon JY. Respiratory syncytial virus disease burden in adults aged 60 years and older in high-income countries: A systematic literature review and meta-analysis. Influenza Other Respi Viruses. 2023;17:e13031. 10.1111/irv.13031

4. Mazur NI, Higgins D, Nunes MC, Melero JA, Langedijk AC, Horsley N, Buchholz UJ, Openshaw PJ, McLellan JS, Englund JA et al. The respiratory syncytial virus vaccine landscape: lessons from the graveyard and promising candidates. Lancet Infect Dis. 2018 Oct;18(10):e295–e311. doi: 10.1016/S1473-3099(18)30292-5 Epub 2018 Jun 18

5. Topalidou X, Kalergis AM, Papazisis G. Respiratory Syncytial Virus Vaccines: A Review of the Candidates and the Approved Vaccines. Pathogens 2023, 12, 1259. 10.3390/pathogens12101259

6. US Food and Drug Administration, Abrysvo® package insert. Available at https://www.fda.gov/vaccines-blood-biologics/abrysvo

7. US Food and Drug Administration, Arexvy® package insert. Available at https://www.fda.gov/vaccines-blood-biologics/arexvy

8. Papi A, Ison MG, Langley JM, Lee D-M, Leroux-Roels I, Martinon-Torres F et al. Respiratory Syncytial Virus Prefusion F Protein Vaccine in Older Adults. N Engl J Med 2023;388:595–608. DOI: 10.1056/NEJMoa2209604

9. Walsh EE, Pérez Marc G, Zareba AM, Falsey AR, Jiang Q, Patton M et al. 2023 Efficacy and Safety of a Bivalent RSV Prefusion F Vaccine in Older Adults. N Engl J Med 2023;388:1465–77. DOI: 10.1056/NEJMoa2213836

10. Wilson E, Goswami J, Baqui AH, Doreski PA, Perez-Marc G, Zaman K, et al. Efficacy and Safety of an mRNA-Based RSV PreF Vaccine in Older Adults. N Engl J Med 2023;389:2233–44.DOI: 10.1056/NEJMoa2307079

11. Wilson E, Goswami J, Doreski PA, Perez-Marc G, Jimenez G, Priddy F, Lin N, Le Cam N, Slobod K, Stoszek SK et al. Efficacy and Safety of mRNA-1345, an RSV Vaccine, in Older Adults: Results through ≥6 Months of Follow-up. RSVVW ReSVINET Conference February 13-16 2024, Mumbai India. Abstract p.88. https://resvinet.org/wp-content/uploads/2024/02/Abstract-Booklet-09Feb24.2.pdf

12. Ison MG, Papi A, Athan A, Feldman RG, Langley JM, Lee DG, Leroux-Roels I, Martinon-Torres F, Schwarz T, van Zyl-Smit RN et al. Efficacy and Safety of Respiratory Syncytial Virus (RSV) Prefusion F Protein Vaccine (RSVPreF3 OA) in Older Adults Over 2 RSV Seasons. Clin Infect Dis. 2024 Jan 22:ciae010. doi: 10.1093/cid/ciae010. Online ahead of print

13. Walsh EE, Falsey A, Patton M, Stacey H, Eiras DP, Jiang Q, Woodside J, Mikati T, Kalinina E, Cooper D et al. Efficacy of a Bivalent RSVpreF Vaccine in Older Adults beyond a First RSV Season. RSVVW ReSVINET Conference February 13–16 2024, Mumbai India.Abstract p.99. https://resvinet.org/wp-content/uploads/2024/02/Abstract-Booklet-09Feb24.2.pdf

14. Falsey AR, Williams K, Gymnopoulou E, Bart S, Ervin J, Bastian AR, Menten J, De Paepe E, Vandenberghe S, Chan EKH et al. Efficacy and safety of an Ad26.RSV.preF- RSV preF protein vaccine in older adults. N Engl J Med. 2023;388:609–620

15. Jenkins, V.A.; Hoet, B., Hochrein, H., De Moerlooze, L. The Quest for a Respiratory Syncytial Virus Vaccine for Older Adults: Thinking beyond the F Protein. Vaccines 2023, 11, 382. 10.3390/vaccines1102038

16. NCT04908683. A Study of an Adenovirus Serotype 26 Pre-fusion Conformation- stabilized F Protein (Ad26. RSV. preF) Based Respiratory Syncytial Virus (RSV) Vaccine in the Prevention of Lower Respiratory Tract Disease in Adults Aged 60 Years and Older (EVERGREEN). https://classic.clinicaltrials.gov/ct2/show/results/NCT04908683?term=vaccine+efficacy+Janssen&cond=RSV&draw=2&rank=1

17. Endt K, W. Y. (2022). A recombinant MVA-Based RSV Vaccine Induces T cell and Antibody Responses that Cooperate in the Protection Against RSV Infection. Front Immunol, 10.3389/fimmu.2022.841471

18. Samy N, Reichhardt D, Schmidt D, Chen LM, Silbernagl G, Vidojkovic S et al. Safety and immunogenicity of novel modified vaccinia Ankara-vectored RSV vaccine: A randomized phase I clinical trial. Vaccine 38 (2020) 2608–2613. 10.1016/j.vaccine.2020.01.055

19. Jordan E, Lawrence SJ, Meyer TPH, Schmidt D, Schultz S, Mueller J et al. Broad Antibody and Cellular Immune Response From a Phase 2 Clinical Trial With a Novel Multivalent Poxvirus-Based Respiratory Syncytial Virus Vaccine. The Journal of Infectious Diseases® 2021;223:1062–72

20. Jordan E, Kabir G, Schultz S, Silbernagl G, Schmidt D, Jenkins VA et al. Reduced Respiratory Syncytial Virus Load, Symptoms, and Infections: A Human Challenge Trial of MVA-BN-RSV Vaccine. The Journal of Infectious Diseases, Volume 228, Issue 8, 15 October 2023, Pages 999–1011

21. World Medical Association. World Medical Association Declaration of Helsinki: ethical principles for medical research involving human subjects. JAMA 2013;310:2191–4. doi:10.1001/jama.2013.281053

22. ICH Guideline for Good Clinical Practice E6 (R2) Step 5. Available at: ICH E6 (R2) Good clinical practice - Scientific guideline | European Medicines Agency (europa.eu)

23. Suter M, Meisinger-Henschel C, Tzatzaris M, et al. Modified vaccinia Ankara strains with identical coding sequences actually represent complex mixtures of viruses that determine the biological properties of each strain. Vaccine 2009; 27:7442–50

24. Falsey AR, Walsh EE, Osborne RH, Vandendijck Y, Ren X, Witek J, Kang D, Chan E, Scott J, Ispas G (2021) Comparative assessment of reported symptoms of influenza, respiratory syncytial virus, and human metapneumovirus infection during hospitalization and post-discharge assessed by Respiratory Intensity and Impact Questionnaire: Influenza and Other Respiratory Viruses 2021

25. US Food and Drug Administration. Guidance for industry: toxicity grading scale for healthy adult and adolescent volunteers enrolled in preventive vaccine clinical trials. 2007 (https://www.fda.gov/media/73679/download)

26. Centers for Disease Control and Prevention. (2023, March). Retrieved from RSV Surveillance: https://www.cdc.gov/surveillance/nrevss/rsv/natl-trend.html

27. RKI SurvStat. (2023, March). Retrieved from Robert Koch Institute: https://survstat.rki.de/Content/Query/Create.aspx

28. Committee for Medicinal Products for Human Use (CHMP). EMA/227054/2023, CHMP assessment report, Arexvy. 26 April 2023

29. US Food and Drug Administration, Abrysvo Cinical Review Memorandum: https://www.fda.gov/vaccines-blood-biologics/abrysvo

30. Fink Al, Klein SL. Sex and gender impact immune responses to vaccines among the elderly. Physiology (Bethesda). 2015 Nov;30(6):408–16. doi: 10.1152/physiol.00035.2015

31. Chambers C, Skowronski DM, Rose C, Serres G, Winter AL, Dickinson JA, Jassem A, Gubbay JB, Fonseca K, Drews SJ, Charest H, Martineau C, Petric M, Krajden M. Should Sex Be Considered an Effect Modifier in the Evaluation of Influenza Vaccine Effectiveness? Open Forum Infect Dis. 2018 Sep 4;5(9):ofy211. doi: 10.1093/ofid/ofy211. PMID: 30263903; PMCID: PMC6143149

32. Ursin RL, Klein SL. Sex Differences in RespiratoryViral Pathogenesis and Treatments. Annu. Rev. Virol. 2021. 8:393–414

33. Dayananda P, Chiu C, Openshaw P.. Controlled Human Infection Challenge Studies with RSV. Curr Top Microbiol Immunol. 2022 Jun 16. doi: 10.1007/82_2022_257. Online ahead of print

34. Salaun B, De Smedt J, Vernhes C, Moureau A, Öner D, Bastian AR, Janssens M, Balla-Jhagjhoorsingh S, Aerssens J, Lambert C, Coenen S, Butler CC, Drysdale SB, Wildenbeest JG, Pollard AJ, Openshaw PJM and Bont L. (2023) T cells, more than antibodies, may prevent symptoms developing from respiratory syncytial virus infections in older adults. Front. Immunol. 14:1260146. doi: 10.3389/fimmu.2023.1260146

35. Walsh EE, Falsey AR, Scott DA, Gurtman A, Zareba AM, Jansen KU et al. A Randomized Phase 1/2 Study of a Respiratory Syncytial Virus Prefusion F Vaccine. The Journal of Infectious Diseases® 2022;225:1357–66

36. Falloon J, Yu J, Esser MT, Villafana T, Yu L, Dubovsky F, Takas T, et al. An Adjuvanted, Postfusion F Protein–Based Vaccine Did Not Prevent Respiratory Syncytial Virus Illness in Older Adults. The Journal of Infectious Diseases® 2017;216:1362–70

37. Novavax. Novavax Announces Topline RSV F Vaccine Data from Two Clinical Trials in Older Adults [Internet]. 2022 [cited 2022Nov10]. Available from: https://ir.novavax.com/2016-09-25-Novavax-Announces-Topline-RSV-F-Vaccine-Data-from-Two-Clinical-Trials-in-Older-Adults

38. US Food and Drug Administration, Jynneos® package insert. Available at https://www.fda.gov/vaccines-blood-biologics/jynneos

39. Nave, L.; Margalit, I.; Tau, N.; Cohen, I.; Yelin, D.; Lienert, F.; Yahav, D. Immunogenicity and Safety of Modified Vaccinia Ankara (MVA) Vaccine—A Systematic Review and Meta-Analysis of Randomized Controlled Trials. Vaccines 2023, 11, 1410. 10.3390/vaccines110914100

